# Higher BMI, but not obesity-related genetic polymorphisms, correlates with reduced structural connectivity of the reward network

**DOI:** 10.1101/2020.05.06.20087577

**Authors:** Frauke Beyer, Rui Zhang, Markus Scholz, Kerstin Wirkner, Markus Loeffler, Michael Stumvoll, Arno Villringer, A. Veronica Witte

**Author notes:** The authors declare no competing financial interests in relation to this work. <u>Corresponding author:</u> A. Veronica Witte, PhD, Department of Neurology, Max-Planck-Institute of Human Cognitive and Brain Sciences, Stephanstr. 1a, 04103 Leipzig.

## Abstract

**Background:** Obesity is of complex origin, involving genetic and neurobehavioral factors. Most consistently, polymorphisms in the fat-and-obesity associated gene (FTO) may increase the risk for developing obesity by modulating dopaminergic signaling in the brain. Dopamine-dependent behaviors, such as reward processing, are crucial for eating behavior and are altered in obesity. Yet, few studies have investigated the association of obesity, related genetic variants and structural connectivity of the dopaminergic reward network.

**Methods:** We analyzed 378 participants (age range: 20 – 59 years, BMI range: 17 – 38 kg/m^2^) of the LIFE-Adult Study. Genotyping for the single nucleotid polymorphisms rs1558902 (FTO) and rs1800497 (near dopamine D2 receptor) was performed on a micro-array. Structural connectivity of the reward network was derived from diffusion-weighted magnetic resonance imaging at 3 Tesla using deterministic tractography of Freesurfer-derived regions of interest. Using graph metrics, we extracted summary measures of clustering coefficient and connectivity strength between frontal and striatal brain regions, normalized for global connectivity. We applied linear regression models to test the association of BMI, risk alleles of both variants and reward network connectivity.

**Results:** Higher BMI was significantly associated with reduced connectivity strength for fractional anisotropy (β= −0.0011, 95%-C.I. [−0.0019, −0.0003], p= 0.0062) and number of streamlines (β = −0.0026, 95%-C.I.:[−0.004,−0.0009], p= 0.0024), but not clustering coefficient. Strongest associations were found for right accumbens, right lateral orbitofrontal cortex and left putamen. As expected, the polymorphism rs1558902 in FTO was associated with higher BMI (F=7.9, p<0.001). None of the genetic variants was associated with reward network structural connectivity.

**Conclusions:** Here, we provide evidence that higher BMI correlates with reduced reward network structural connectivity. This result is in line with previous findings of obesity-related decline in white matter microstructure. We did not find any association of variants in FTO or near DRD2 receptor and reward network structural connectivity, indicating that the genetic influence of these variants is small or non-existent. Future research should investigate the behavioral implications of structural connectivity differences in the fronto-striatal network and incorporate larger sample sizes with longitudinal designs in order to gain further insight into the genetic determinants of obesity in the brain.

## Introduction

Obesity or excess body weight is the result of an imbalance in energy intake and expenditure and is now mainly considered a neurobehavioral disorder which involves homeostatic brain regions as well as regions engaged in salience and reward processing (1). Although increasing obesity rates in western societies may be primarily driven by the availability of high-fat diets and a sedentary lifestyle (2), the risk of developing obesity is under strong (polygenetic) control, with heritability estimates of 50%-70% (3, 4). Yet, it is poorly understood which individual (genetic) predispositions determine the vulnerability to those environmental influences and how they lead to excessive weight gain and obesity.

Single nucleotid polymorphisms in the fat and obesity related gene (FTO) are the most common genetic variants associated with obesity (5). Yet, little is known about the mechanisms underlying this association (6). Variation in FTO may contribute to the risk for obesity by modulating feeding behavior rather than energy expenditure (7). Here, one possible mechanism is that polymorphisms in FTO influence dopamine signaling between the Nucleus accumbens (Nacc) and frontal brain regions, either directly or by interaction with dopamine-related variants (8). Consequently, behavioral changes in reward processing, learning and impulsivity might lead to differences in eating behavior, and ultimately, weight gain (9–12). Functional neuroimaging studies have provided evidence for a main effect of FTO on the neural response to food cues, and reported that this association depended on fasting state and ghrelin signaling (13–17). Further studies have shown that FTO interacted with the Taq1A polymorphism, located near the D2 dopamine receptor (DRD2), to modulate reward learning and neural activity in response to food cues – both key functions of the Nacc (1820).

Besides functional activation, the strength of structural connections between the Nacc and frontal brain regions plays an important role for dopamine-dependent behavior (21, 22). Van Schouwenburg and colleagues showed that the effects of a dopaminergic drug depended on fronto-striatal structural connectivity (23). Previously, individual effects of dopaminergic drugs have been related to baseline working memory capacity, which presumably reflects baseline dopamine levels (24). Along these lines, a DRD2 variant (rs6277 C-allele), previously linked to reduced striatal binding potential, was associated with increased fronto-stri-atal structural connectivity (25). This variant is in linkage disequilibrium with the Taq-1A (D’=0.66) (26).

Regarding the structural connectivity of the reward network in obesity, studies showed contradicting results, with both higher and lower structural connectivity between striatum and frontal cortex associated with weight status (27–29). For other white matter tracts, studies have consistently reported associations of higher BMI and lower WM microstructure, possibly mediated by the negative metabolic impact of obesity (30, 31). Recently, a variant in FTO was shown to interact with weight status on fronto-striatal structural connectivity, indicating that some of the mixed results might be due to genetic variability or differences in tonic dopamine levels (32, 33).

In summary, while there is consistent evidence of obesity-related differences in global white matter microstructure, less is known about the association of BMI and reward network white matter structure, and the impact of obesity-related genotypes on this trait. In this study, we aimed to investigate the association of structural reward network connectivity, obesity and genetic variations linked to obesity in a well-characterized population-based sample. Based on previous studies, we hypothesized that a higher BMI would relate to lower reward network connectivity (27). More exploratory, we investigated whether FTO and Taq1A polymorphisms had interactive or independent effects on structural connectivity within the reward network (19, 32). This may help to better understand the genetic and neurobehavioral background of obesity.

## Methods

### Participants

All participants took part in the MRI assessment of the LIFE-Adult-Study (n ~ 2600)(34). The study was approved by the institutional ethics board of the Medical Faculty of the University of Leipzig and conducted according to the declaration of Helsinki. Besides age and sex stratification, participants were randomly selected from the registry of the city of Leipzig and gave written informed consent. We included participants aged in between 20 and 59 years (n ~ 800) with complete information on BMI and genotyping (n ~ 400). Further, we excluded participants with stroke, cancer treatment in the last 12 months, neuroradiological findings of brain pathology, epilepsy, multiple sclerosis, Parkinson’s disease or intake of centrally active medication (n = 17).

Due to incomplete fat suppression during the diffusion-weighted imaging (DWI) acquisition, we observed an artifactual rim in the parietal lobe of the brain. As this artifact was difficult to detect in an automated fashion, we manually rated all scans into three categories dependent on the number and severity of affected slices, i.e. “no/very mild”, “moderate”, and “severe”. We restricted our analysis to those scans that were rated as “no/very mild” and “moderate”. Two participants were excluded due to outlying values in structural connectivity (see Connectome reconstruction from DWI-data). This led to a total sample of 378 healthy volunteers eligible for analysis.

### Genotyping

Genomic DNA was extracted from EDTA treated blood samples using the Autopure LS instrument (Qiagen, Hilden). Genotyping of FTO polymorphism rs1558902 and rs1800497 (Taq1A), a polymorphism near dopamine D2 receptor, was performed based on using the Affymetrix Axiom Technology with custom option (Axiom-CADLIFE). Individuals were imputed at the 1000Genomes reference phase 1, release 3 (http://mathgen.stats.ox.ac.uk/impute/data_download_1000G_phase1_integrated.html) using SHAPEIT v2 and IMPUTE 2.3.0. Details of the measurement and quality control can be found elsewhere (35). The risk allele of the polymorphism rs1558902 on FTO is the A allele and genotype frequencies for rs1558902 were TT n=126, AT n=186, AA n=66 (minor allele frequency = 0.42, test for Hardy-Weinberg-Equilibrium Chi^2^=.03, df=1, n.s.). We considered three groups of risk carriers for the analysis (0: TT, 1: AT, 2: AA).

For the rs1800497 polymorphism the genotype frequencies were TT n=270, AT n=100, AA n=8 (minor allele frequency = 0.15, test for Hardy-Weinberg-Equilibrium Chi^2^=.13, df=1, n.s.). We grouped together carriers of at least one risk allele (A).

### Anthropometric measurement

Anthropometric measurements were conducted by trained study personal. Body weight was measured with scale SECA 701, height was measured with height rod SECA 220 (SECA Gmbh & Co. KG). Body mass index (BMI) was calculated as the weight in kilograms divided by the square height in meters (kg/m^2^).

### Possible confounders

We performed sensitivity analysis of our results by considering different confounders. Head motion is an important confounder in neuroimaging studies, not only affecting measures of functional connectivity, but also measures of brain structure. Since we previously noticed a strong collinearity between BMI and head motion during resting-state, we adjusted for head motion in our analysis of structural connectivity (36). Therefore, we used the six rotation and translation parameters returned by eddy_correct during preprocessing to calculate mean framewise displacement as described by (37). Mean FD was used to adjust for head motion during the DWI scan.

Smoking status and education were derived from self-report questionnaires. Smoking status was available for N=364 participants in three levels (0 = never smoker, 1 = previous smoker, 2 = current smoker). Education was given in 3 levels (1 = left school without degree or finished secondary school after 9 years, 2 = finished secondary school after 10 years, 3 = finished secondary school after 12 or 13 years) and was available for N=376 participants. Depression scores were derived from the CES-D questionnaire as previously described and were available in n=355 participants (38). We log-transformed mean FD and CES-D score prior to the analysis because of the skewed distribution of these scores.

### MR data acquisition

Brain imaging was performed on a 3T Verio Scanner (Siemens, Erlangen) with a 32 channel head coil. T1-weighted images were acquired using generalized autocalibrating partially parallel acquisition imaging technique (39) and the Alzheimer’s Disease Neuroimaging Initiative standard protocol with the following parameters: inversion time, 900 ms; repetition time (TR), 2300 ms; echo time, 2.98 ms; flip angle, 9°; band width, 240 Hz/pixel; image matrix, 256 × 240; 176 partitions; field of view, 256 × 240 × 176 mm3; sagittal orientation; voxel size, 1 × 1 × 1 mm3; no interpolation.

Diffusion-weighted imaging (DWI) was performed using a double spin echo sequence with the following parameters: TR, 13.8 s; TE, 100 ms; image matrix, 128 × 128 × 72; field of view, 220 × 220 × 123 mm3, voxel size of 1.7 mm isotropic, max b-value = 1000 s/mm2, 60 directions. T2*-weighted functional images were acquired using an echo-planar-imaging sequence with the following parameters: repetition time, 2s; echo time, 30 ms; flip angle, 90°; image matrix, 64 × 64; 30 slices; field of view, 192 × 192 × 144 mm3, voxel size of 3 mm × 3 mm, slice thickness of 4 mm, slice gap of 0.8 mm; 300 volumes; total acquisition time, 10:04 minutes.

### T1-data processing

Cortical reconstruction and volumetric segmentation of T1-weighted MR images was performed with the Freesurfer image analysis suite (version 5.3.0) (40, 41). Regions of interest from subcortical segmentation and cortical Desikan-Killiany parcellation were used as nodes in connectome reconstruction. Volumetric measures of reward network regions (bilateral lateral and medial orbitofrontal cortex, caudate, putamen and accumbens) were averaged across hemispheres for analysis.

### Connectome reconstruction from DWI data

Connectome reconstruction was performed during the 10kin1day-workshop, a collaborative science event where over 10000 DWI datasets were processed. More information and summary data can be found online (42). Preprocessing included correction for susceptibility and eddy currents using FSL’s tool eddy-correct. Freesurfer’s standard Desikan-Killiany parcellation was used to select 68 cortical and 14 subcortical regions for connectome reconstruction. A diffusion tensor was fitted to each voxel in a white matter mask using robust tensor fitting and deterministic pathway tractography was applied to construct white matter fiber tracts. Tracts were started in each voxel and then followed the main diffusion direction using the fiber assignment by continuous tracking (FACT) algorithm. Stopping criteria were FA-value below 0.1, crossing of the brain mask or a fiber turn of more than 45 degrees. Weighted, unsigned and undirected connectivity matrices were constructed for each subject by using 82 (sub)cortical regions as nodes and connectivity weights between the regions as edges. Two types of connectivity weights were assessed 1) total number of connecting streamlines touching both regions (NOS) and 2) mean FA across voxels included in these streamlines (FA).

Reward connectivity networks were reconstructed by using bilateral lateral and medial or-bitofrontal cortex, caudate, putamen and accumbens as nodes and their respective structural (NOS and FA-strength) connectivity strength as edges (27). Data quality assurance was done according to the 10kin1day-workshop guidelines by identifying subjects with outlying values in mean NOS or mean FA connectivity of existing connections or outlying values in prevalence of existing or non-existing connections. Outliers were defined as values higher than (3rd quartile + 2*interquartile range (IQR)) or lower than (1st quartile - 2*interquartile range (IQR)). This led to the exclusion of two participants with bad data quality (excessive head motion and large ventricles).

### Reward network analyses

Reward connectivity networks were reconstructed by using bilateral lateral and medial or-bitofrontal cortex, caudate, putamen and accumbens as nodes and their respective structural (NOS and FA-strength) connectivity strengths as edges. Graph metrics were calculated using the Matlab-based Brain Connectivity Toolbox. For each participant, whole brain connectivity was described using mean connectivity strength (CS) and mean clustering coefficient (CC) across all nodes with mean FA and NOS as structural weights. The NOS structural connectivity weights were normalized to [0, 1] prior to calculating the weighted clustering coefficient. Graph metrics were calculated in the reward network accordingly and normalized by the corresponding whole brain value (27), resulting in the following reward network (REW) metrics: FA CS, FA CC, NOS CS, and NOS CC.

For sensitivity analysis, we assessed normalized node-wise connectivity strength and clustering coefficient for the ten nodes (bilateral lateral and medial orbitofrontal cortex, caudate, putamen and accumbens) in the network.

### Statistical analysis

We used univariate linear models in R version 3.3.2 to investigate the association of the BMI, genetic variants and measures of reward network (REW) structural connectivity (43). We use F-tests and associated p-values from R’s anova to compare full versus null models. First, we tested the association of BMI and the four reward network measures, adjusting for age and sex (Model 1: REW metric ~ Age + sex + BMI). If there was a significant association of BMI, tested by comparing this full model against a null model including only age and sex (Bonferroni-corrected alpha=0.0125), we additionally adjusted for average head motion during the DWI scan, smoking status, depression and education (Model 2: REW metric ~ Age + sex + head motion + smoking status + depression + education + BMI).

We further investigated the node-wise connectivity if the edge type (i.e. FA or NOS) reached network-wide significance. Thereby we aimed to identify nodes that contributed most to the result. Here, we fitted Model 2 (node-wise REW metric ~ Age + sex + head motion + smoking status + depression + education + BMI) and used Bonferroni-corrected alpha=0.005 (accounting for 10 regions of interest tested).

In the genetic analysis, we first investigated whether the polymorphisms rs1558902 and rs180047 explained variance in BMI. We compared a full model including the interaction of rs1558902 (0/1/2 risk allele groups) with rs180047 (0/1 or 2 risk allele groups) (Model 3: BMI ~ Age + sex + FTO* Taq1A) against a null model without the interaction and main effects (Model 4: BMI ~ Age + sex + FTO + Taq1A). Without a significant interaction (Chi-Square test of Model 3 vs. 4: p<0.05), we report only main effects of model 4.

Then, we tested whether the polymorphisms were associated with reward network connectivity, either independently or interacting with each other. Again, we first tested the interaction effect of the two variants (Model 5: REW metric ~ Age + sex + FTO*Taq1A) by comparing it to a null model without the interaction. If there was no indication for an interaction (p> 0.05), we checked for independent effects of the two genotypes separately (Model 6: REW metric ~ Age + sex + FTO + Taq1A), again by comparing against a null model including Age, sex. If we found a significant effect (p<0.05) for any of the two variants, we additionally included head motion, smoking status, depression, education as covariates. The data used in this study is available upon request from the authors.

## Results

In total, we analyzed 378 participants (55% women), for descriptive statistics see **Tab. 1**.

**Table 1.**
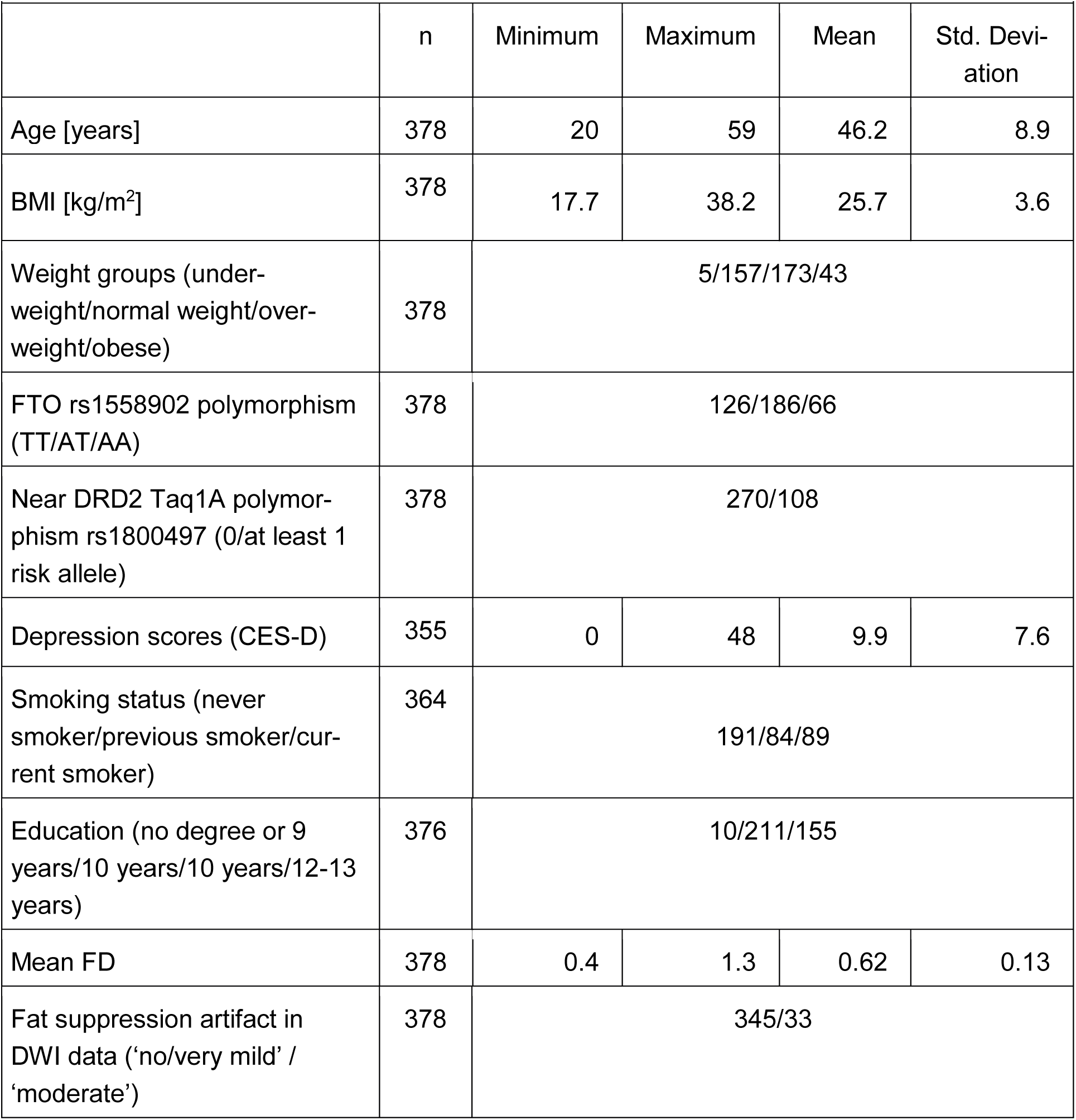
Demographic characteristics of the sample (n=378, sex distribution: 170 men and 208 women). Weight groups according to WHO definition. BMI, body mass index; CES-D, Center for Epidemiologic Studies Depression Scale; mean FD, mean framewise displace-ment during diffusion-weighted imaging.

|  |  |  |  |  |  |
| --- | --- | --- | --- | --- | --- |
|  | n | Minimum | Maximum | Mean | Std. Deviation |
| Age [years] | 378 | 20 | 59 | 46.2 | 8.9 |
| BMI [kg/m<sup>2</sup>] | 378 | 17.7 | 38.2 | 25.7 | 3.6 |
| Weight groups (under-weight/normal weight/over-weight/obese) | 378 | 5/157/173/43 |  |  |  |
| FTO rs1558902 polymorphism (TT/AT/AA) | 378 | 126/186/66 |  |  |  |
| Near DRD2 Taq1A polymorphism rs1800497 (0/at least 1 risk allele) | 378 | 270/108 |  |  |  |
| Depression scores (CES-D) | 355 | 0 | 48 | 9.9 | 7.6 |
| Smoking status (never smoker/previous smoker/current smoker) | 364 | 191/84/89 |  |  |  |
| Education (no degree or 9 years/10 years/10 years/12-13 years) | 376 | 10/211/155 |  |  |  |
| Mean FD | 378 | 0.4 | 1.3 | 0.62 | 0.13 |

|  |  |  |
| --- | --- | --- |
| Fat suppression artifact in DWI data ('no/very mild' / 'moderate') | 378 | 345/33 |

### Association of BMI and reward network metrics

In the main linear regression analysis, reconstruction of structural connectivity showed that a higher BMI was associated with lower strength of both FA and NOS edge measures in the reward network, after adjusting for age and sex (Bonferroni corrected, all p < 0.0062; model 1, **Fig. 1**; **Tab. 2**). Results remained stable when additionally correcting for potential confounding effects of head motion, smoking status, depression score and education (model 2). The CC for both FA and NOS were not associated with BMI.

**Fig. 1:**
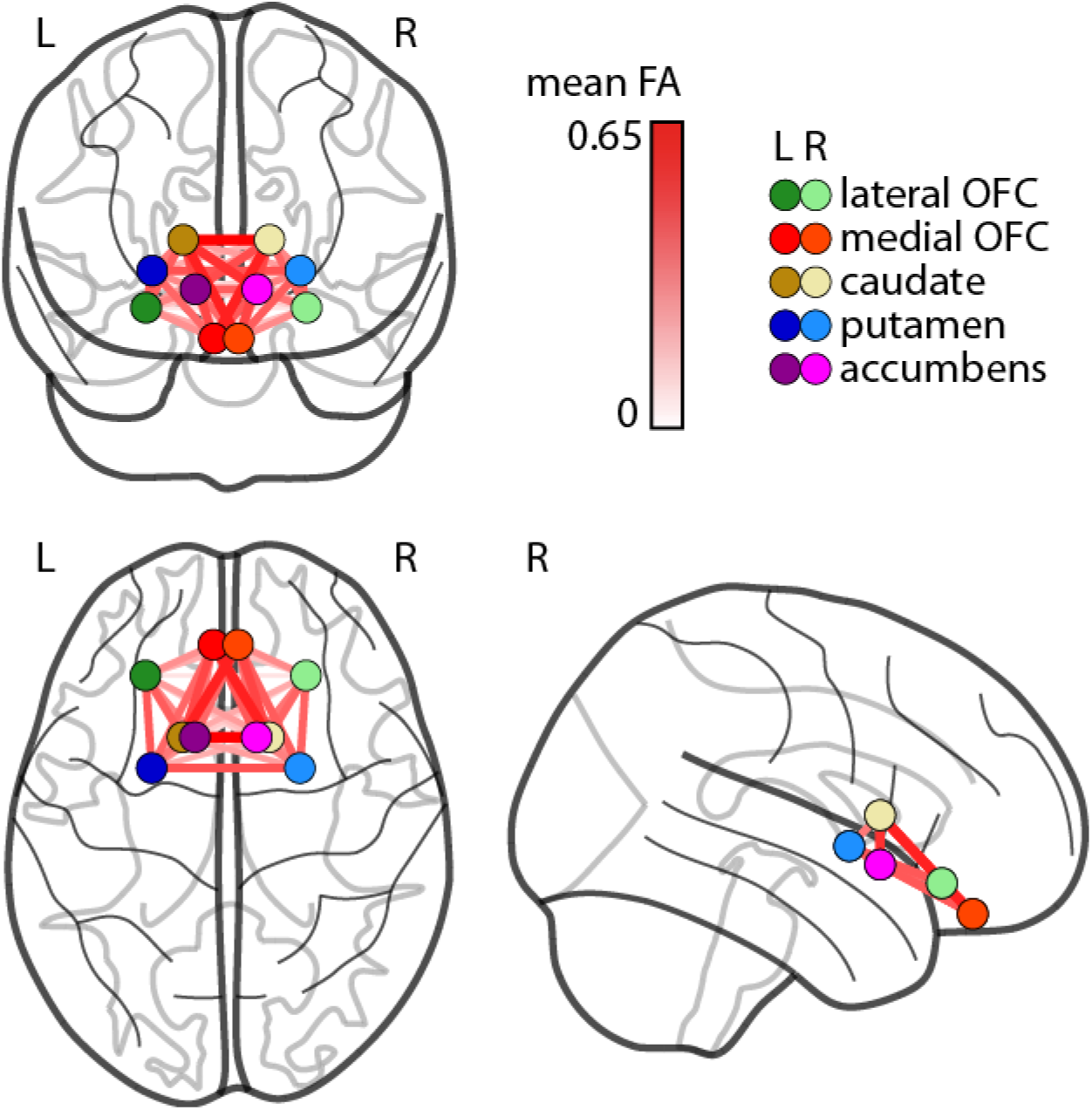
Structural connectivity measured using mean fractional anisotropy (FA) between hubs of the reward network. OFC, orbitofrontal cortex; L, left hemisphere; R, right hemisphere.

**Table 2:** Results from the linear regression analysis of BMI and reward network structural connectivity strength (CS) and clustering coefficient (CC) with edge weights fractional anisotropy (FA) and number of streamlines (NOS). Given values are adjusted R^2^, β, 95% - confidence interval (C.I.) of β & p for the effect of BMI (Model 1 & 2).

|  |  | FA CS | FA CC | NOS CS | NOS CC |
| --- | --- | --- | --- | --- | --- |
| Model 1 | Adj. $R^2$ | 0.12 | 0.11 | 0.12 | 0.002 |
| | $\beta$ | -0.00108 | -0.024 | -0.0026 | 0.25 |
|  | C.I. | [-0.0019, -0.0003] | [-0.046, -0.0032] | [-0.004, -0.0009] | [-0.13, 0.63] |
|  | p | 0.0062 | 0.024 | 0.0024 | 0.19 |
| Model 2 | Adj. $R^2$ | 0.11 | - | 0.09 | - |
| | $\beta$ | -0.00102 | - | -0.0026 | - |
|  | C.I. | [-0.0019, -0.0002] | - | [-0.004, -0.0007] | - |
|  | p | 0.02 | - | 0.0073 | - |

In addition, within-network node-wise measures showed that for FA, lower CS of the bilateral Ncl accumbens and putamen, as well as right lateral OFC, were significantly associated with higher BMI (**Tab. 3**). For NOS, lower CS of the left putamen and left medial OFC was significantly associated with higher BMI (**Tab. 3**).

**Table 3:** Results from the linear regression analysis of BMI and node-wise connectivity strength (CS) with edge weights fractional anisotropy (FA) and number of streamlines (NOS). Given are adjusted R^2^, β, 95% - confidence interval (C.I.) of β and p-value for the effect of BMI (Model 2, adjusted for age, sex, depression, smoking status, education, head motion during scan). Bonferroni-corrected, significant p-values are highlighted in **bold**.

| FA CS | hemi-sphere | Adj. $R^2$ | $\beta$ | C.I. | p |
| --- | --- | --- | --- | --- | --- |
| Nucleus accumbens | Right | 0.09 | -4.3 e-04 | [-7.2 e-04; -1.3 e-04] | **0.0043** |
|  | Left | 0.08 | -3.8 e-04 | [-7.2 e-04; -4.4 e-05] | 0.026 |
| Caudate | Right | 0.03 | 1.8 e-04 | [-4.8 e-05; 4.1 e-04] | 0.12 |
|  | Left | 0 | -3.7 e-05 | [-2.4 e-04; 1.6 e-04] | 0.72 |
| Putamen | Right | 0.10 | -2.4 e-04 | [-4.4 e-04; -4.3 e-05] | 0.017 |
|  | Left | 0.07 | -3.4 e-04 | [-5.5 e-04; -1.3 e-04] | **0.0018** |
| Medial OFC | Right | 0.09 | -7.9 e-05 | [-2.7 e-04; 1.1 e-04] | 0.43 |
|  | Left | 0.08 | -1.8 e-04 | [-3.8 e-04; 2.1 e-05] | 0.08 |
| Lateral OFC | Right | 0.07 | -4.6 e-04 | [-7.9 e-04; -1.3 e-04] | **0.0056** |
|  | Left | 0.07 | -8.0 e-05 | [-4.1 e-04; 2.5 e-04] | 0.63 |
| **NOS CS** |  |  |  |  |  |
| Nucleus accumbens | Right | 0.05 | -5.2 e-04 | [-1.0 e-03; -4.2 e-05] | 0.03 |
|  | Left | 0.06 | -4.2 e-04 | [-8.6 e-04; 2.5 e-06] | 0.05 |
| Caudate | Right | 0.05 | -3.7 e-05 | [-5.1 e-04; 5.8 e-04] | 0.89 |
|  | Left | 0 | -4.0 e-04 | [-9.7 e-04; 1.6 e-04] | 0.17 |
| Putamen | Right | 0.07 | -7.2 e-04 | [-1.5 e-03; 3.9 e-05] | 0.06 |
|  | Left | 0.08 | -1.4 e-03 | [-2.2 e-03; -5.3 e-04] | **0.0015** |
| Medial OFC | Right | 0.03 | -3.1 e-04 | [-9.2 e-04; 2.9 e-04] | 0.32 |
|  | Left | 0.05 | -7.0 e-04 | [-1.3 e-03; -8.7 e-05] | 0.025 |
| Lateral OFC | Right | 0.04 | -3.6 e-04 | [-8.5 e-04; 1.4 e-04] | 0.16 |
|  | Left | 0.05 | -4.1 e-04 | [-8.6 e-04; 4.7 e-05] | 0.08 |

### Association of genotypes and BMI

There was a trend for an interaction effect of rs1558902 and Taq1A on BMI (adjusted for age and sex, F=2.9, p=0.056) seen as a less pronounced effect of Taq1A in carriers of one FTO risk allele. In the following, we report the model without interaction. There was a significant increase in BMI with increasing number of rs1558902 risk alleles (adjusted for age and sex, F=7.9, T_AAvsTT_=2.9, T_AAvsTT_=3.7, all p<0.001). Carriers of at least one risk allele in rs180047 had a trend towards lower BMI compared to non-risk allele carriers (adjusted for age and sex, F=3.6, T_AT/AAvsTT_=-0.73, p=0.057). (**Fig. 2**).

**Fig. 2:**
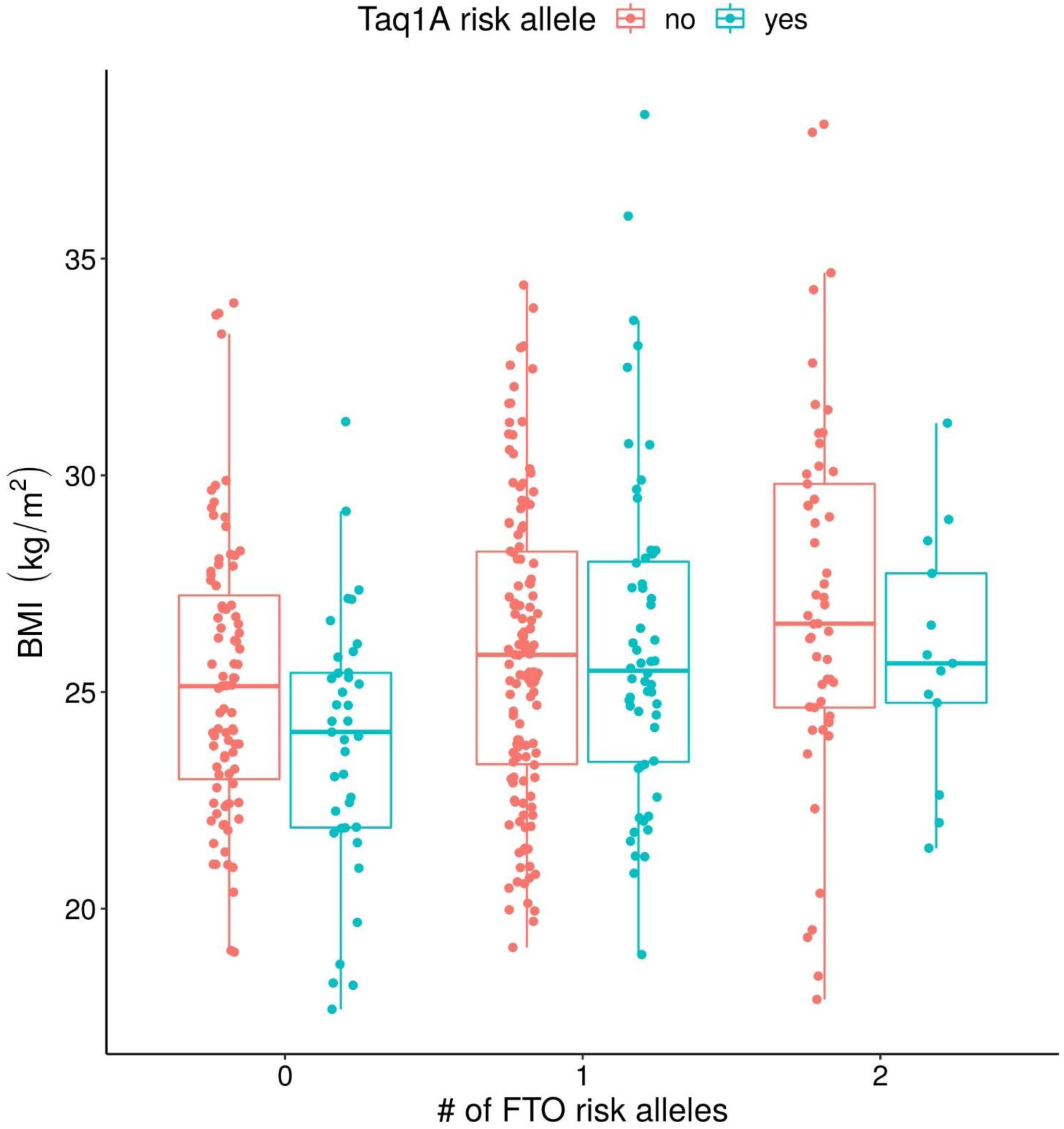
Body mass index (BMI) depends on FTO- and Taq1A genotypes. Individual data points are shown for carriers of 0, 1 or 2 FTO risk alleles (x-axis) with no or at least one Taq1A risk allele (red/blue, respectively). Bars within boxes give medians, surrounding boxes show the interquartile range, vertical lines indicate 1.5 times the interquartile range above/below the upper/lower quartile, respectively.

We did not observe a significant interaction of FTO and Taq1A genotypes on reward network structural connectivity when adjusting for age and sex. Also, there were no significant main effects of these polymorphisms (see Table 4 for more details).

**Table 4:**
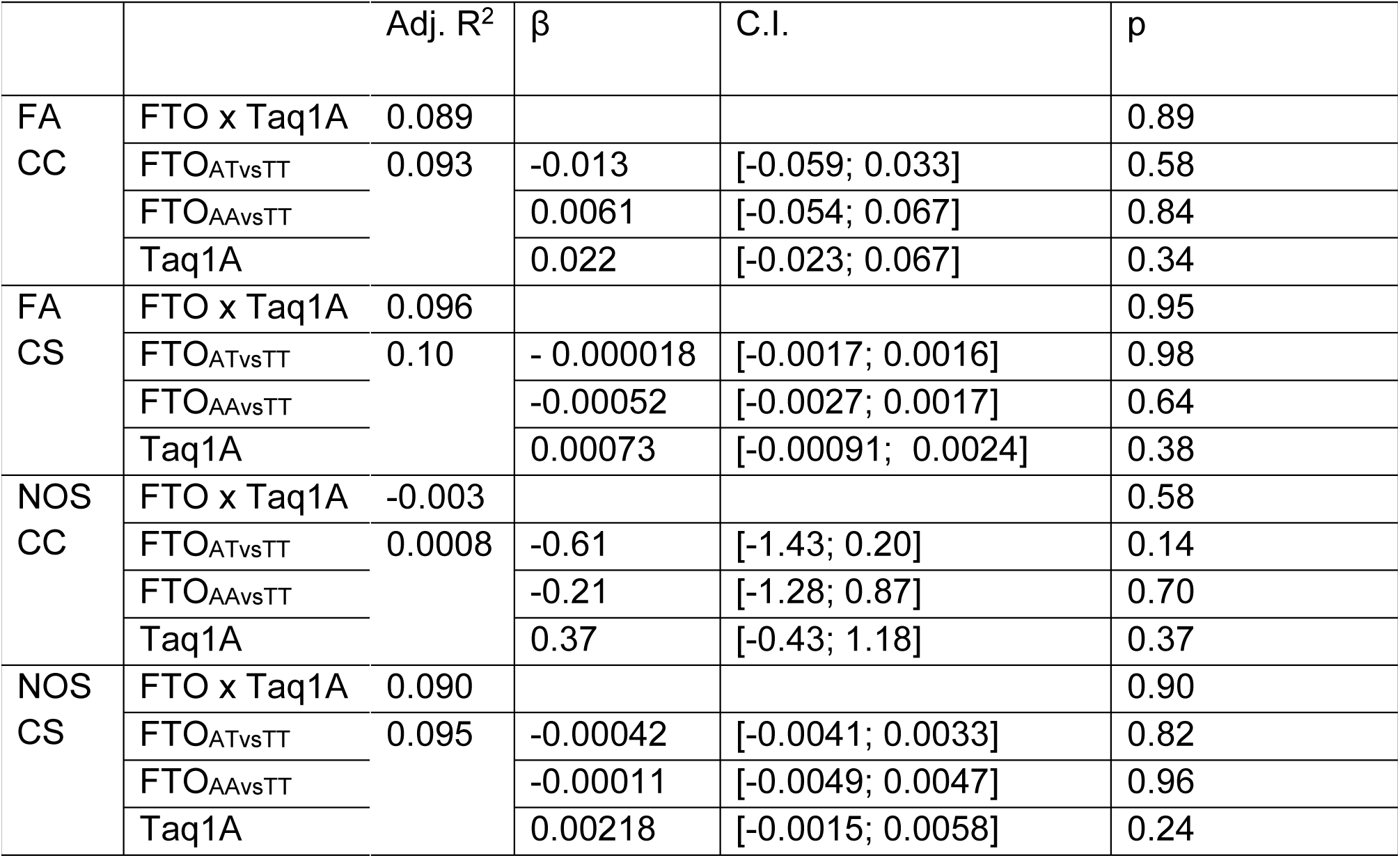
Results from linear regression analysis of BMI, genetic variants and reward network structural connectivity. For the FTO × Taq1A interaction, the adjusted R^2^ value of the regression model and the p-value of the interaction, derived from comparison with a null model including age and sex, are shown. For individual genetic effects the adjusted R^2^ value of the regression model, and the regression coefficients β for Taq1A, FTO_ATvsTT_ (0 vs. 1 risk allele) and FTO_AAvsTT_ (0 vs. 2 risk alleles), 95% - confidence intervals (C.I.) of β coefficients and p-value of the coefficients are given. FA CC: clustering coefficient of the reward network based on fractional anisotropy, FA CS: connectivity strength of the reward network based on fractional anisotropy, NOS CC: clustering coefficient of the reward network based on number of streamlines, NOS CS: connectivity strength of the reward network based on number of streamlines.

## Discussion

In this cross-sectional analysis in young to middle-aged adults, a higher BMI was significantly associated with lower structural connectivity of the reward network. More specifically, higher BMI predicted less connectivity strength (CS) for edge weights in the Nacc, putamen and orbitofrontal cortex, even if adjusting for age, sex, education and other conditions such as depressive symptoms. In addition, while obesity-related SNPs in the FTO gene significantly predicted higher BMI, we did not observe genetic associations with measures of structural connectivity in the reward network.

Our results of lower reward network connectivity with higher BMI is in line with a previous study that examined obesity-related differences in measures of white matter structural coherence. Using the same graph-based approach, Marqués-Iturria et al. (2015) reported lower connectivity strength and network clustering of the striatum and orbitofrontal cortex in 31 obese participants compared to 32 lean controls (27). In contrast, (44) reported higher structural connectivity of the bilateral putamen in obese and overweight participants compared to normal weight controls. This study also investigated sex-by-weight status interactions and found that among individuals with overweight and obesity women had stronger connectivity of reward network regions than men. While we chose the same definition of structural connectivity as Marqués-Itturia, Gupta et al. derived their connectivity values from the number of connections and used different graph metrics. This might be an explanation for the diverging results. Yet, analyses in larger samples support our finding and showed that higher BMI was associated with lower FA within white matter tracts throughout the brain, including connections between reward network regions (30, 31, 45). Longitudinal studies suggest that, at least in older adults, these differences may reflect damage due to the metabolic consequences of vascular risk factors (46, 47). Regardless of its origin, reduced structural connectivity in the reward network might underlie certain behavioral traits observed in obesity, such as aberrant processing of food-related reward stimuli (12, 48)

Considering genetic effects, we replicated the known association of FTO risk alleles and higher BMI, while carrying the Taq1A risk allele was not associated with higher BMI in our cohort. We did not find an association between the genetic variants in FTO and near DRD2 on reward network structural connectivity, in contrast to previous results which proposed that white matter microstructure may be genetically linked to obesity: In a large cohort of Mexican-American families, BMI and white matter microstructure in different fiber tracts were indeed genetically correlated (49). Dennis and colleagues reported that among 15 obesity-associated genetic variants, a variant in NEGR1 had the strongest effect on fractional anisotropy in the corona radiata in a sample of ~500 young adults (50). Further, cumulative effects of all considered obesity-associated genotypes predicted white matter microstructure in arcuate fasciculus, corpus callosum, fornix, and inferior frontal-occipital fasciculus.

Other studies have provide evidence for a link between genetic variants in dopamine D2/D3 receptors (DRD2), reward processing and obesity (51, 52), even though the role of variation in DRD2 in obesity is still debated (53, 54). For example, both the Taq1A variant in the DRD2 receptor and a genetic sum score reflecting higher dopamine signaling capacity predicted future increases in BMI (55–57). Regarding structural connectivity, both FTO and a genetic variant in strong linkage disequilibrium with Taq1A correlated with structural connectivity of striatal-frontal tracts in two studies of small to moderate sample size (25, 32). Yet, we could not replicate these findings. There may be several reasons for this. First, the true effect of these variants is still probably small, thus we may have been underpowered to show it. Alternatively, the influence of environment (e.g. metabolic conditions associated with obesity) or gene-environment interactions might blur the link between genetic factors and brain structure in our cohort.

In summary, future studies incorporating larger samples sizes and longitudinal designs need to further address the underlying (epi-)genetics of reward structural connectivity and obesity.

### Limitations

In this study, we did not investigate the causal relationship between BMI and structural connectivity of the reward network. Furthermore, our sample was not rich in obese participants, which might limit the power of our analyses. Although our sample size was adequate for an imaging study, we might be underpowered to detect subtle effects of single polymorphisms on brain connectivity. Finally, we did not assess epigenetic factors in this design. Strengths of our study include the well-characterized cohort that enabled us to adjust for important confounders in our analysis, and the high-resolution DWI protocol.

### Conclusions

We here provide evidence that higher BMI is associated with reduced reward network structural connectivity. We did not find any contribution of variants in FTO or near DRD2 receptor gene to reward network structural connectivity, indicating that the genetic influence of these variants is small or non-existent. Future research should investigate the behavioral implications of structural connectivity differences in the fronto-striatal network and incorporate larger sample sizes with longitudinal designs in order to gain further insight into the genetic determinants of obesity in the brain.

## Data Availability

The data that these results are based on is available from the authors upon request.

## Acknowledgments

We would like to thank all participants and staff of the LIFE-Adult study. This work was supported by grants of the European Union, the European Regional Development Fund, the Free State of Saxony within the framework of the excellence initiative, the LIFE-Leipzig Research Center for Civilization Diseases, University of Leipzig [project numbers: 713-419 241202, 14505/2470, 14575/2470] and by grants of the German Research Foundation, contract grant number CRC 1052 “Obesity mechanisms” Project A1, A. Villringer/M. Stumvoll, WI 3342/3-1, A.V. Witte and by the Max Planck Society.

